# Validation of Angular Indication Measurement (AIM) Stereoacuity

**DOI:** 10.1101/2025.04.23.25325959

**Authors:** Sonisha Neupane, Jan Skerswetat, Peter J. Bex

**Affiliations:** Northeastern University, College of Science, Department of Psychology, Boston, Massachusetts, United States of America; University of California, Irvine, School of Medicine, Department of Ophthalmology, 850 Health Sciences Road, Irvine, California, 92697, United States of America

**Keywords:** Depth perception, stereoacuity, response-adaptive psychophysics, binocular vision

## Abstract

**Background:** Stereopsis is a critical visual function, however clinical stereotests are time-consuming, coarse in resolution, suffer low test-retest repeatability, and poor agreement with other tests. We developed AIM Stereoacuity to address these limitations and asked whether it could deliver reliable, efficient, and precise stereo-thresholds across stimulus types and disparity signs.

**Methods:** Observers reported the orientation of 5×1.25° bar defined by disparity of random dots embedded in a 6° diameter circular cell, presented in a 4*4 grid. Bar disparity was scaled from ± 2σ relative to a threshold and slope-estimate, initially set by the experimenter and adaptively updated. Orientation report errors (indicated vs. actual bar-orientation) were fit with a cumulative Gaussian function to derive stereo-thresholds. Twenty-one normally-sighted observers were tested with red-blue anaglyphs in crossed and uncrossed disparity signs across 4 element-types (8.5arcmin broadband dots, or band-pass difference of Gaussians with peak Spatial-Frequency (SF) of 2, 4, or 6 c/°). We analyzed stereoacuities, test durations, and the test-retest repeatability.

**Results:** Across SFs and observers, test duration for a chart were 36 and 40 secs for measuring crossed and uncrossed disparity, respectively. There was no effect of disparity sign or SF (Kruskal-Wallis; p>0.05). Median log stereo-thresholds averaged across all SFs were 1.90 and 1.84 log arcsec for crossed and uncrossed disparities, respectively. Crossed and uncrossed disparities were moderately correlated across SFs(r=0.44 to 0.79; median=0.54). Test-retest biases were 0.01 arcsec (p=0.45) and 0.10 arcsec (p= 0.001) for crossed and uncrossed disparities, respectively.

**Conclusions:** The results for the response-adaptive, self-administered AIM Stereoacuity method showed no significant stereo-thresholds differences between broad- and narrow-band stimuli. The test delivers repeatable results for crossed disparity in approximately 80 seconds.

## Introduction

Stereoacuity, the ability to perceive depth through binocular vision, plays a fundamental role in various aspects of daily life, from precise motor tasks to spatial awareness in near visual space (1). Current clinical stereo-tests typically measure stereopsis using booklet tests. These tests have several problems such as poor agreement between different tests (2–4), low repeatability (5), coarse stereoacuity resolution, memorization artifacts, low sensitivity to changes, and monocular artifacts (6, 7), depending on the test. Furthermore, these tests require a trained administrator and also are limited to a fixed, narrow range of stimulus properties (8) such as broad-band Wirth circles, broad band random dot noise, or single disparity polarity, i.e. either crossed or uncrossed.

We recently developed the Angular Indication Measurement (AIM) paradigm, which is a generalizable, rapid, self-administrable, and response-adaptive paradigm to quantitatively assess a range of visual functions through an orientation report error paradigm (9, 10). We recently adapted a stereoacuity paradigm for AIM that eliminates or reduces the above-mentioned problems by utilizing a continuous report paradigm and validated the approach (9). In this study, we use AIM Stereoacuity to investigate the impact of stimulus type and depth-direction on stereoacuity, which could provide valuable insights into its potential applications in clinical settings and perceptual research.

Stereoacuity is influenced by a range of stimulus properties, such as luminance (11), contrast (12, 13), spatial frequency (14), and direction i.e., crossed/uncrossed disparity (15, 16) or orientation (17).

Spatial frequency (SF) plays a role in stereoacuity sensitivity. Stimuli with peak SF around 5 c/° have been shown to elicit maximum sensitivity to stereoscopic disparity (14, 18). The rate of change in stereo-thresholds across retinal eccentricity is also influenced by SF (19) and the loss of stereopsis due to factors like refractive error or amblyopia may vary across different SFs (20–22). This suggests that individuals with various visual disorders—such as amblyopia, strabismus, or macular degeneration— may exhibit different patterns of stereo-loss depending on the spatial frequency properties of the stimulus used.

The direction of disparity (crossed/near or uncrossed/far) and various stimulus properties also influence stereoacuity (15, 16, 23–26). However, most current tests measure only one direction (typically crossed disparity) and use a limited range of stimuli, often differing between tests. A study has shown that detection thresholds for crossed disparities are lower than those for uncrossed disparities, which may be attributed to different binocular mechanisms (15). Additionally, the ability to process crossed disparity develops earlier than the ability to process uncrossed disparity in infants (15).

Although the orientation of disparity has not been extensively studied, it is known to impact stereoacuity. Blake et al. observed that stereoacuity was highest with vertical rods and progressively decreased as the angle approached horizontal (17).

The current study had four goals: to measure the test durations for each chart and stimulus type, to compare the stereoacuities for different spatial frequencies and disparity directions, to validate the AIM Stereoacuity’s test-retest repeatability, and to investigate the influence of stimulus orientation on stereoacuity for each stimulus condition in stereo-typical participants.

## Methods

The study was approved by the Institutional Review Board at Northeastern University (14-09-16) and followed the guidelines of the Declaration of Helsinki. The recruitment period was from September 2022 to October 2022. Informed written consent was obtained from all participants prior to the start of the experiment. The participants were undergraduate students who completed the study for course credit.

21 normally-sighted observers with an age range from 18-22 participated in the study. All participants wore their habitual glasses or contact lenses, if any, and had visual acuity of 20/20-20/30 with correction as verified via ETDRS visual acuity test prior to the start of the main experiments.

### Stimuli and Procedure

Stimuli were created with MATLAB software (MathWorks, Version 2021b and 2023b) in combination with Psychtoolbox (27–29). Stimuli were presented on a gamma-corrected 32” 4K LG monitor with maximum luminance of 250/m^2^ and screen resolution of 3840 x 2160 and 60Hz at 80 cm viewing distance. A chin rest ensured the viewing distance was consistent. Red-blue anaglyph glasses were used to present the stimuli to each eye dichoptically. Crossed and uncrossed stereo-depth was achieved using horizontal disparity offsets between the left and right eye.

AIM is a generalizable (10, 30–32), self-administered, and response-adaptive method which presents a sequence of charts that consist of a grid of cells, each containing a target. The current study validated the stereoacuity version of the AIM platform, called *AIM Stereoacuity*. Three charts i.e., one initial chart and two response-adaptive subsequent charts were deployed for each stimulus condition. Each chart contained a 4*4 grid of 6° diameter circular cells, each surrounded by a white indication ring of 0.1° line width. Each cell contained 100 random dot stimuli presented dichoptically to generate horizontal stereo-disparity (see Figure 1A for example charts). Four stimulus conditions were investigated in a randomized order, 8.5 arcmin diameter broadband dots, or band-pass difference of Gaussians (DoGs) with SF_peak_ at 2, 4 or 6c/°. Stereoscopic disparity was introduced to dots within a 5° x 1.2° rectangular bar oriented randomly within the dot stimuli backgrounds. This test employed a red-blue anaglyph display and glasses to present stimuli in both crossed and uncrossed disparity signs. The stereoscopic disparities of the first chart ranged from 1° to 1’, in 16 evenly spaced log steps, which spans the performance range of a typical population (33), thus AIM Stereoacuity’s initial chart includes easy- and difficult-to-detect stimuli for participants with or without mild stereo-impairment. A stereoscopic disparity threshold was estimated based on these initial responses using a psychometric function (see equation 1). The mid-point (δ_τ_) and slope (σ) of this function served as a personalized estimate for performance that was used to scale the disparities of bars in each subsequent chart in 16 evenly spaced log steps that ranged from difficult (threshold -2σ) to easy (threshold +2σ) to detect. Participants had unlimited time to report the orientation of the depth-defined bars via mouse click on the indication ring around each cell, guessing if necessary. Participants could click on either end of the perceived bar. Their reported orientation was shown by two black (≈0 cd/m²) feedback marks, and they could refine their response with unlimited additional clicks. After indicating the orientation for all cells, participants clicked on a ’Next’ icon at the right side of the screen to move to the next chart. Orientation report errors i.e., the difference between indicated and actual bar orientation of the disparity-defined bar as a function of horizontal disparity were fit with a cumulative Gaussian function to determine stereo-thresholds.

**Figure 1).**
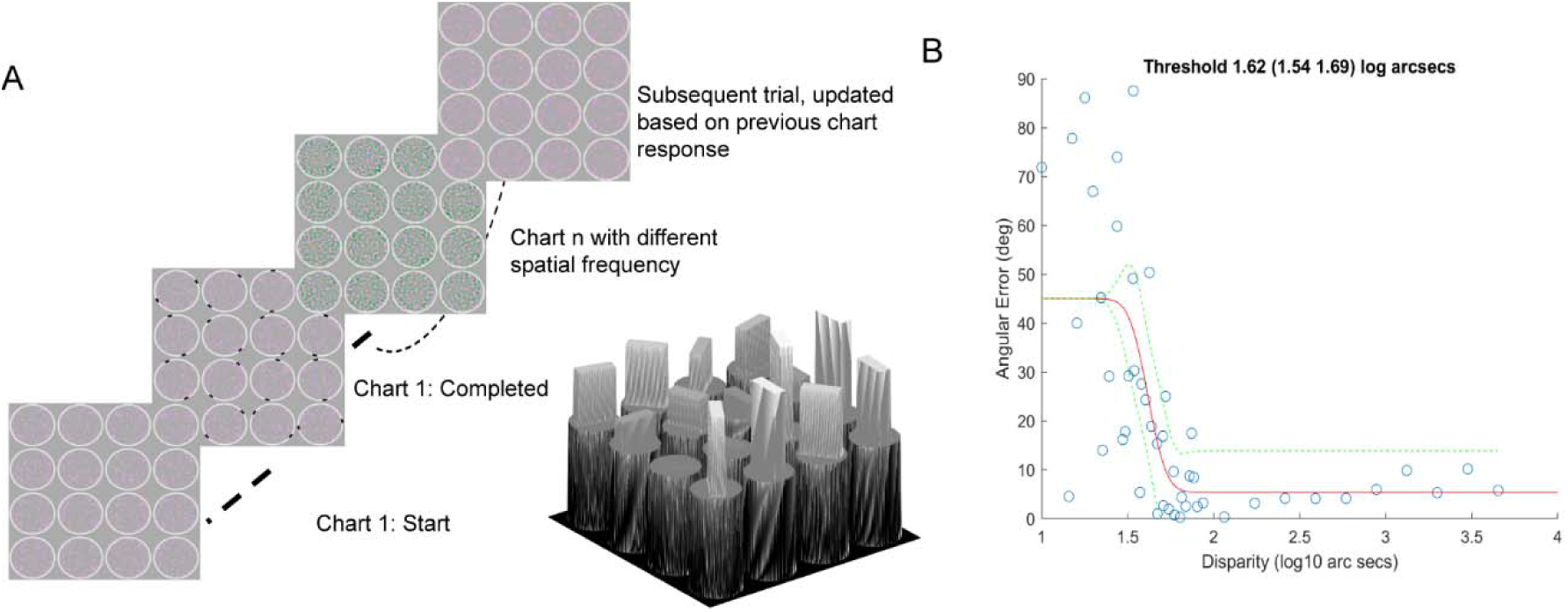
AIM Stereoacuity’s stimulus for dot elements with broadband spatial frequency. The range of disparities presented on the first chart was defined by the experimenter (1° to 1’ in even log-spaced steps) and on subsequent charts was adaptively calculated based on the threshold and slope of each participant’s responses to previous charts. The illustration in grey visualizes the varying degrees of depth due to the varying degrees of horizontal disparity. B) Orientation report error (y axis) as a function of the stereoscopic disparity of the bar (x axis) for a typical naïve participant. Orientation errors (blue circles) for each of the 48 indicated cells as a function of horizontal disparity were fit with a cumulative Gaussian function (equation 1, red curve, ±95% CI green dashed lines) to derive the stereo-threshold.

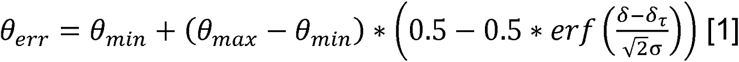

Where θ*_err_* is orientation error, θ*_min_* is the minimum report error for a highly visible target, θ*_max_* is the maximum mean error for guessing (here 45°), δ is stimulus stereoscopic disparity, δ_τ_ is threshold disparity and σ is the slope.

The threshold is calculated as the midpoint between the minimum angular error (θ*_min_*) and the maximum mean error of 45°, corresponding to random responses. All tests were completed twice with a short break between tests, the test order for the different element conditions was randomized between participants.

## Data analysis

Nonparametric Kruskal-Wallis, Wilcoxon signed-rank, and Brown-Forsythe tests were used as the data were not normally distributed. The Matlab function *corrplot* was used to compare linear correlations across tests and stimuli conditions using Kendall rank correlation coefficient. Bland-Altman plots (34, 35) were used to analyze the repeatability of the tests. Matlab *tic* and *toc* commands were used to measure each chart and the total duration of a single stimulus condition.

## Results

### Experiment Duration and Stereoacuity

Figure 2A shows the test durations for a single chart for crossed and uncrossed disparities across 2 runs for 3 charts for 4 stimulus conditions i.e., 24 charts for each participant as swarm plots that are superimposed by boxplots. The test durations from 2 runs were statistically different for both crossed (z(74245)=-6.50, p <0.01; Wilcoxon signed-rank test) and uncrossed disparities (z(75574)=-7.31, p<0.01; Wilcoxon signed-rank test). The test duration were statistically different (z(240569)= -2.96, p<0.01; Wilcoxon signed-rank test) for crossed and uncrossed disparities and the variability was higher for uncrossed disparities (median: 39.8 sec, inter-quartile range (IQR): [26.2 sec]), compared to the crossed condition (median: 35.7 sec, IQR= 20.3 sec).

**Figure 2:**
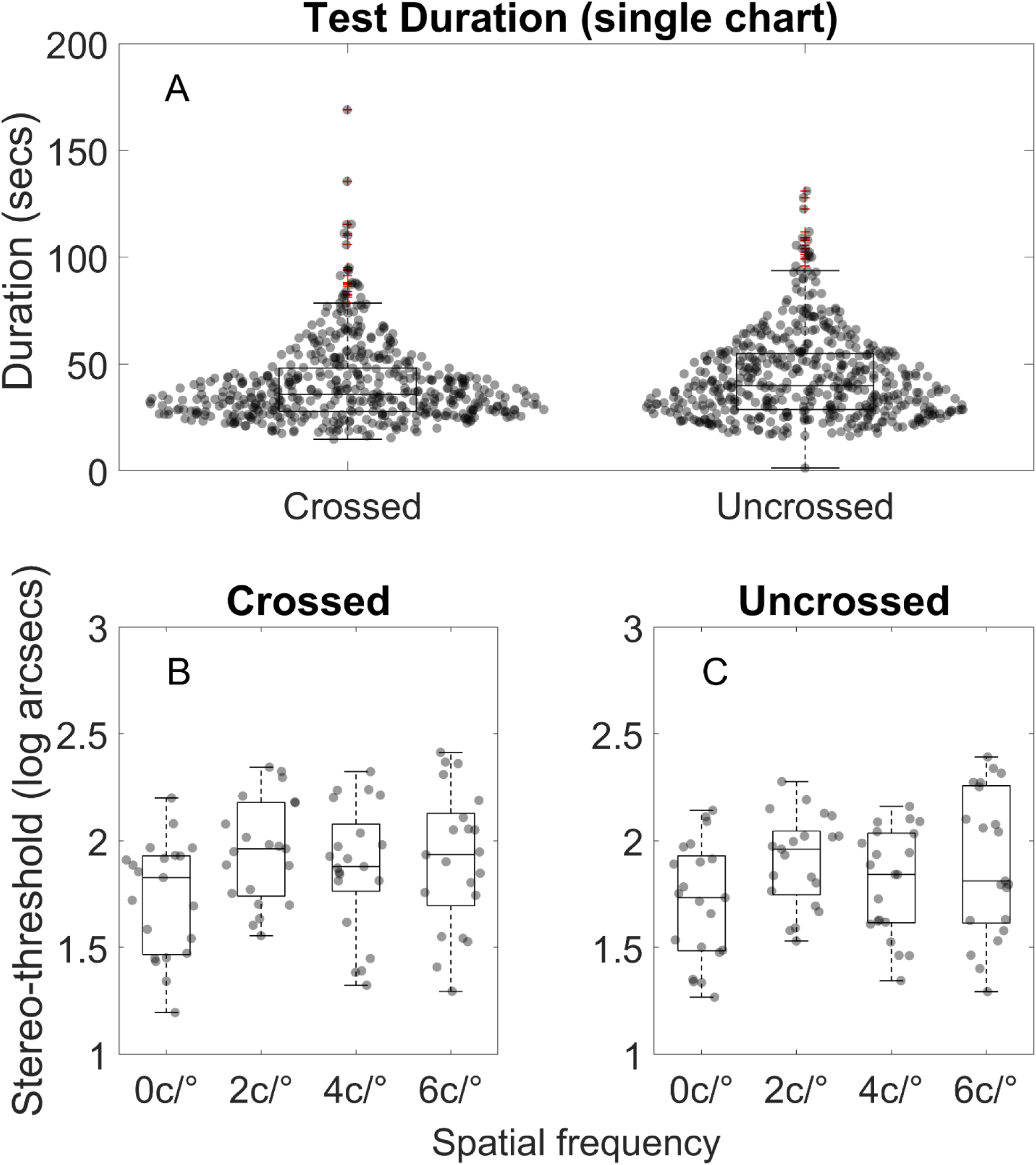
A) Time required to complete a single Stereoacuity charts for crossed and uncrossed disparities. 2B-C) Stereoacuities measured with AIM Stereoacuity for broadband (0c/°) or band-pass (2, 4 or 6 c/°) elements. Datapoints show the results for individual participants averaged across the first and second test for crossed (B) and uncrossed (C) sign. Horizontal lines show the median, boxes show the interquartile range and whiskers show 95% intervals.

Figure 2B and 2C shows the threshold for crossed and uncrossed disparities for stimulus with broadband (0 c/°) or band-pass (2, 4 or 6 c/°) elements. Thresholds for the 2 runs were not statistically different for crossed (z(7139)=0.13, p=0.90; Wilcoxon signed-rank test or uncrossed disparities (z(7671)=1.82, p=0.07; Wilcoxon signed-rank test and were therefore averaged. The disparities with broadband or band pass elements were not statistically different for either crossed (H(3, 80)=6.15, p=0.1045; Kruskal Wallis test) or uncrossed disparities (H(3, 80)=6.03, p=0.1099; Kruskal Wallis test). The median of AIM Stereoacuity’s thresholds for crossed (1.89 log arcsec, [IQR= 0.39 logarcsec]) and uncrossed (1.84 log arcsec, [IQR=0.44 log arcsecs]) disparities were also not statistically significant (z(7189)=0.70, p=0.49; Wilcoxon signed-rank test).

## Correlation results

Figure 3 shows the Kendall’s rank correlation coefficient between crossed and uncrossed disparities for the broadband (0c/°) or band-pass (2, 4 or 6 c/°) elements. All correlations were significantly different from zero (p<0.05).

**Figure 3:**
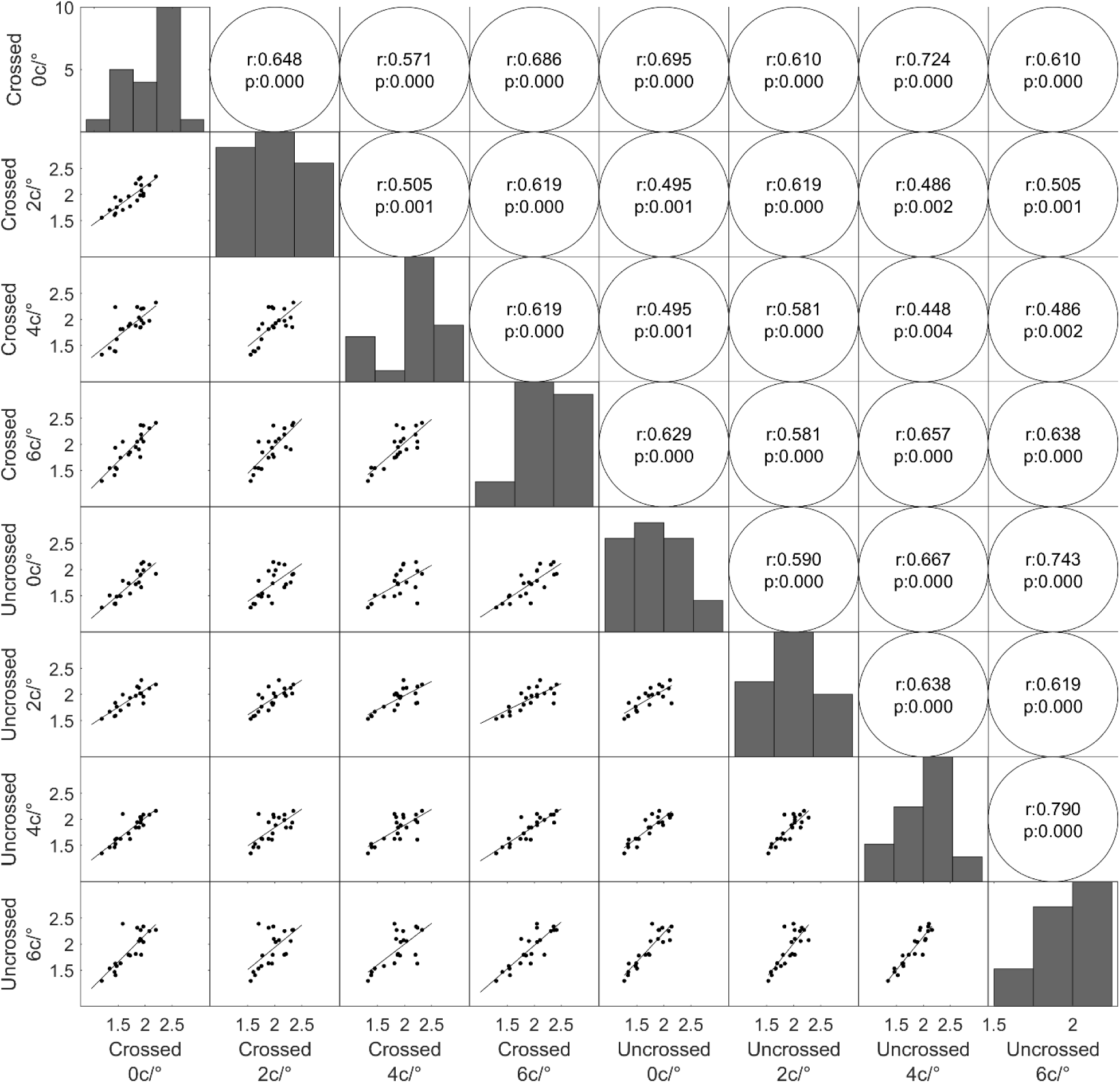
Correlation between crossed (C) and uncrossed (U) stereoacuities (log arcsec) for broadband (0) and SF bandpass (2, 4 or 6 c/°) elements. The dots show data for individual observers, the line shows regression. The bar charts in the diagonal show the frequency distribution for each SF condition. The Kendall’s rank correlation coefficients and p values for each condition are shown in the circles.

## Repeatability results

Figure 4 A and B show the Bland-Altman analyses for repeatability between two completions of the same test from the participants for the crossed and uncrossed disparities. The mean bias for crossed disparity was not significant (mean bias= 0.01 log arcsec; p = 0.45) whereas it was significant for uncrossed (0.10 log arcsec; p = 0.001). Similarly, the proportional bias was also significant for uncrossed disparity (p=0.001) but not for crossed disparities (p=0.43). The coefficient of repeatability of the uncrossed condition (±0.59 log arcsec) was approximately twice as large as that of the crossed condition (±0.34 log arcsec).

**Figure 4:**
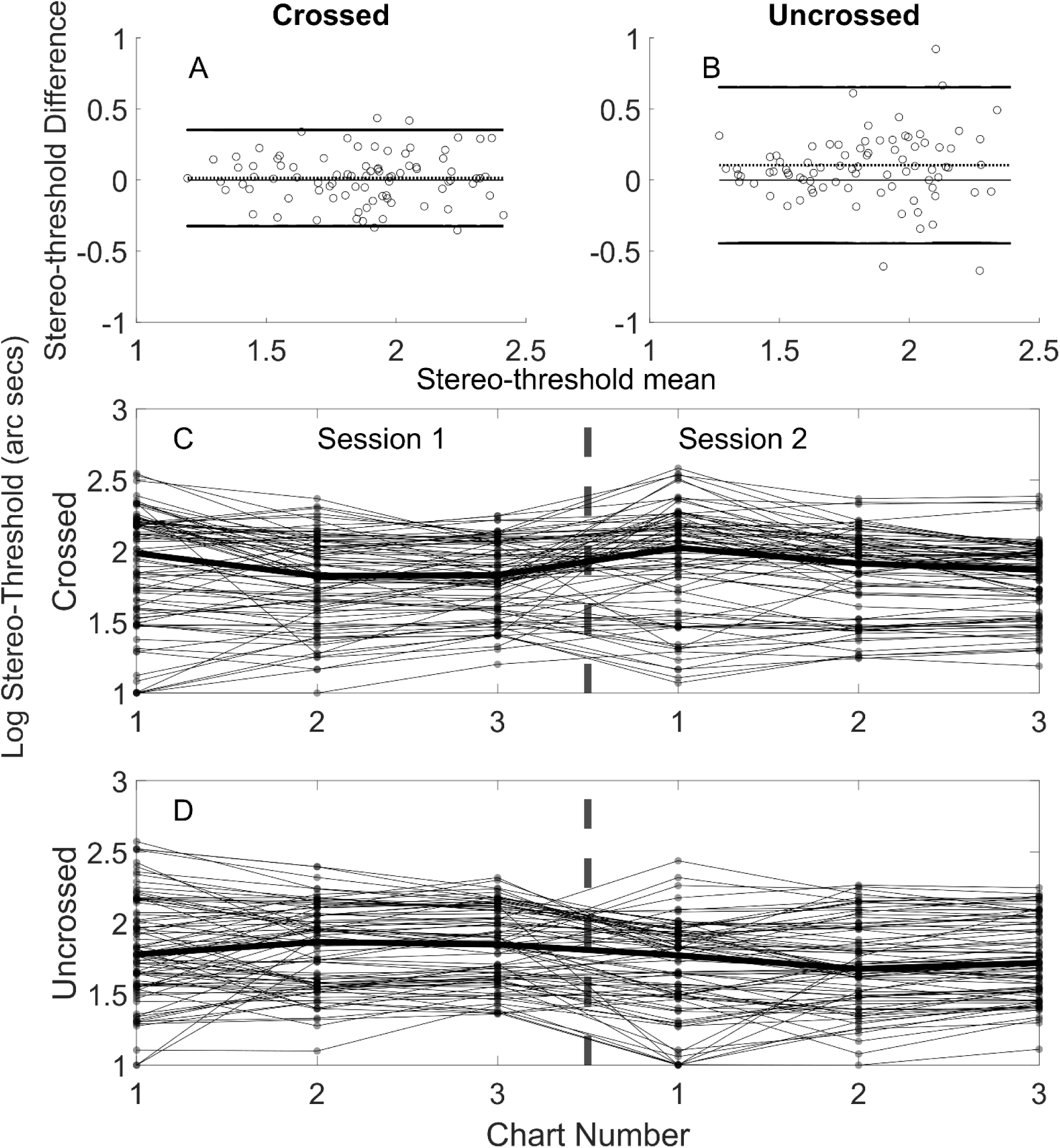
Bland-Altman plots to assess the repeatability between AIM Stereoacuity tests from 2 runs for crossed (A) and uncrossed (B) disparity stimuli. All participants test-retest difference in log stereo-thresholds are shown as circles. The black solid lines indicate the zero difference (as reference) and the upper and lower limits of agreement, the black dashed lines show the mean bias. Figure 4C & D: Stereo-threshold estimates following each chart in the first and second session for crossed and uncrossed disparities. Each line shows individual participants, the bold lines indicated the median for each chart number.

Figure 4 C and D shows estimates of stereo-threshold for crossed and uncrossed disparity as a function of chart number in the first and second session. There is more variability with the first chart in each session, then variability decreases with the addition of each new chart (p=0; Brown-Forsythe Test). For crossed disparities, the stereo-threshold after chart 1 was statistically different than after chart 2 and 3 (H(2, 501)=14.52, p=0.0007; Kruskal Wallis test), whereas stereo-thresholds did not significantly differ across charts for uncrossed disparities (H(2, 501)=0.97, p=0.6142; Kruskal Wallis test).

### Stimulus orientation bias

We also investigated whether there was any systematic orientation report bias. Figure 5 A and D show histograms of orientation report error as a function of target orientation for A) crossed and B) uncrossed stimuli. Data have been combined across all participants, repeats and SF condition into 5° bins. The power spectrum for orientation report errors shows a peak at 4 cycles for crossed (B) and 2 cycles per 360° for uncrossed stimuli (E). Figure C and F show the mean error for target orientation and the concentration parameter (kappa) of von Mises distribution estimates for (C) crossed and (F) uncrossed disparities. Sinewave with 2 cycles and 1 cycle per 180° were fitted over the data respectively.

**Figure 5:**
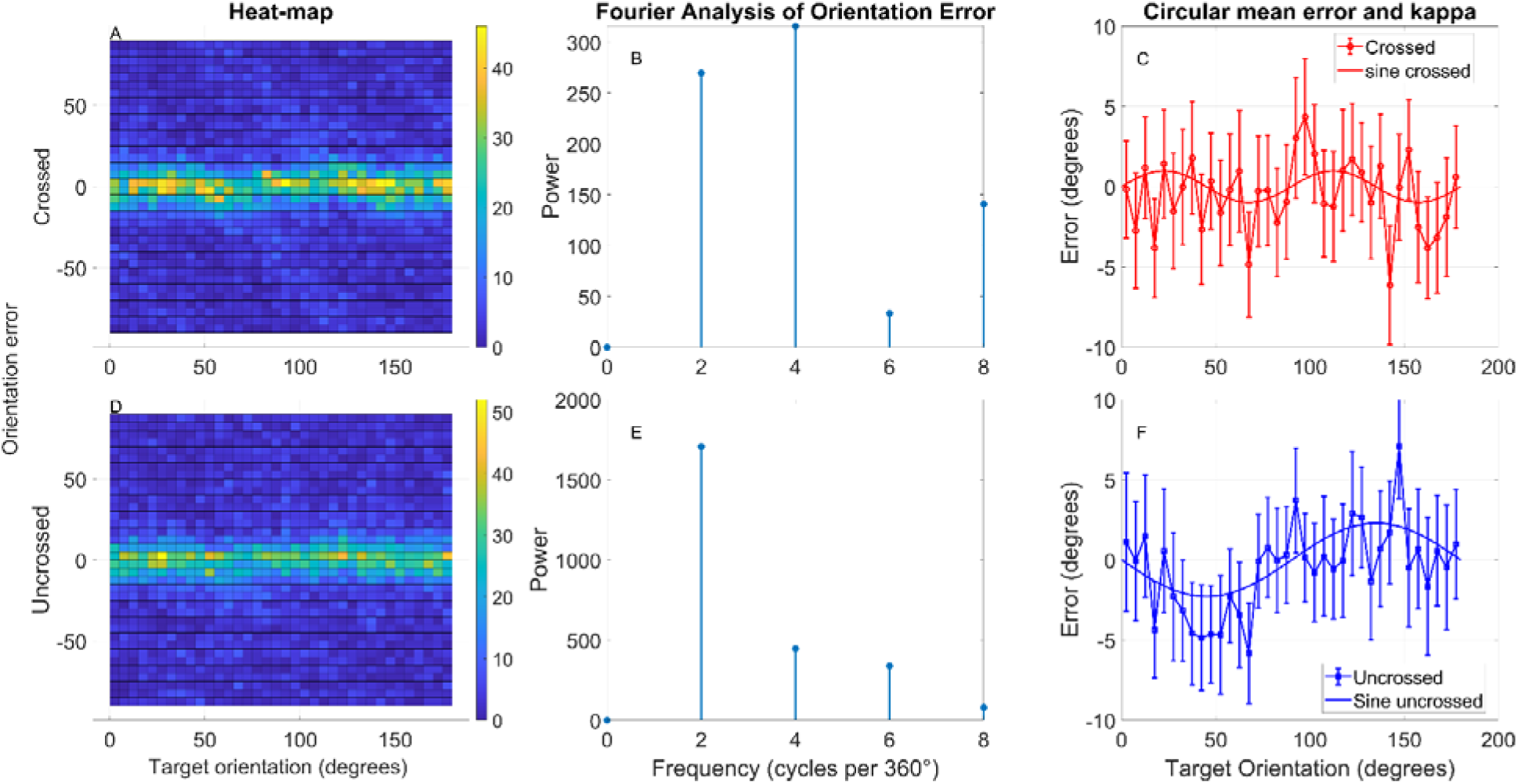
A) Orientation report bias as a function of target orientation for crossed stimuli. The x axis shows the orientation of the target bar (0° = horizontal), the y axis shows a histogram of report errors. The bin size was 5°, the colorbar indicates the frequency of error occurrences. B) Fourier analysis of orientation error. C) Mean error and kappa for crossed stimuli. D-F show similar figures for uncrossed stimuli.

## Discussion

Our results demonstrate that AIM Stereoacuity is capable of rapidly and precisely measuring stereo-thresholds. The average duration for measuring stereo-thresholds for a single chart for uncrossed and crossed disparities was 38 seconds. This finding suggests that AIM Stereoacuity offers an efficient and accurate self-administered method for assessing depth perception, with the ability to achieve results in a relatively short time frame.

One surprising result in our study was the lack of a significant difference in stereo-thresholds between crossed and uncrossed disparities, as well as between different spatial frequencies. Previous studies have consistently shown differences in thresholds for crossed and uncrossed disparities (16, 24–26, 36), as well as variations in stereoacuity based on spatial frequency (14, 18). The absence of these expected differences in our findings may be attributed to the differences in methodology used to determine the stereo-threshold. Our paradigm employed a constant number and density of stimulus elements (dots or DoGs) for all conditions, whereas others have employed sinusoidal variations in depth, in which the density of disparity signals covaries with spatial frequency, which may have influenced the outcome. Additionally, variations in participant responses, including individual differences in binocular function or sensitivity, could have contributed to the lack of significant differences observed.

The correlation between stereo-thresholds for different spatial frequencies and disparity directions ranged from 0.44 to 0.79 (Figure 3), and all correlations were statistically significant. This finding suggests a strong relationship between the stereo-thresholds for both crossed and uncrossed disparities, as well as between the different stimulus structures used. The results indicate that the AIM Stereoacuity is effective in estimating stereoacuities for a broad range of stimulus conditions and provides consistent measurements across various types of disparity, which also identifies redundancies that may reduce test time for screening purposes.

We examined whether the orientation of the target systematically affected the reported direction. As expected for an adaptive paradigm, most errors were within a range of ±10 degrees and were random beyond this range (Figure 5). There were, however, small (<2°) but systematic biases in report error that depended on the target orientation and differed between crossed and uncrossed disparities. For crossed disparities, responses were slightly biased *away* from the cardinal orientations (0° and 90°). This is similar to response repulsion effects frequently observed in orientation paradigms, for review and discussion of whether this is a perceptual or a decision effect, see (37). For uncrossed disparities, however, responses were slightly biased *towards* 0° and 180°. This effect is surprising and inconsistent with other orientation report biases, however, it suggests that biases may differ between crossed and uncrossed disparities and therefore that the biases originate from different sensory processes for crossed and uncrossed disparities, rather than decision or motor processes. The biases are small and would increase the random error parameter (θ*_min_*) of the psychometric function without affecting the threshold estimate, thus would not affect the reliability of the AIM Stereoacuity. These findings show that the test can effectively account for different angles of disparity without significant biases in the results.

Repeatability analyses, plotted using Bland-Altman plots (34) showed a systematic learning effects for uncrossed disparities but not for crossed disparities (Figure 4). The analysis coefficient of repeatabilities for each sign revealed that uncrossed disparities showed more test-retest variance compared to the crossed condition. Furthermore, when comparing the stereo-thresholds obtained with two versus three charts, we found no significant difference for both crossed and uncrossed disparities (Figure 4). Taken together, these results suggest that using two charts in the AIM Stereoacuity provides reliable and accurate stereoacuity measurements in around 80 seconds.

Despite the promising results, there are some limitations to this study that should be considered. First, the test was only conducted with a typical population of young adults, and while the method is likely to be effective for individuals with binocular impairments, this was not directly tested in the present study. Future research will validate AIM Stereoacuity in populations with conditions such as amblyopia, strabismus, or other binocular vision disorders. Additionally, the study sample was restricted to young adults, and the performance in other age groups—such as children and the elderly—was not evaluated. Given that the administration of stereoacuity tests may present unique challenges in these populations, further studies involving children and elderly individuals are necessary to determine the broader applicability of the AIM Stereoacuity across the lifespan.

### Conclusions

The current study validated the self-administered, response-adaptive AIM Stereoacuity and included an assessment of stereo-thresholds for different stimulus properties, test-retest repeatability, testing time, and orientation report-bias analysis in normally-sighted observers. An investigation of target orientation and report error showed small but systematic biases that differed between crossed and uncrossed disparities. The per-chart and the test-retest analyses revealed that only the crossed disparity sign condition generated repeatable and unbiased results with decreasing result variability after at least one adaptive step. The time analysis indicated that AIM Stereoacuity for two charts can therefore be completed in about 80 seconds. Lastly, we found no significant effect for stimulus disparity or spatial composition of the target stimulus. Future research will investigate whether these stimulus properties of AIM Stereoacuity are able to detect spatially-selective vision loss in clinical cohorts such as patients with amblyopia.

## Data Availability

All data produced in the present study are available upon reasonable request to the authors.

## Acknowledgements

Portions of this study have been presented at the Vision Science Society Conference 2022 and Vision Science Society conference 2023.

## Author Contribution Statement

SN was responsible for project administration, methodology, extracting and analyzing data, interpreting results, writing the initial draft and reviewing and editing the draft. JS was responsible for conceptualization, methodology, analyzing and interpreting results and reviewing and editing of the draft. PJB was responsible for conceptualization, funding acquisition, managing resources, project administration, interpretation of data, supervision, reviewing and editing of the draft.

